# Physical inactivity among corporate bank workers in Accra, Ghana: Implications for health promotion

**DOI:** 10.1101/2022.11.08.22282093

**Authors:** George B. Nketiah, Kwasi Odoi-Agyarko, Tom A Ndanu, Frank E. A. Hayford, Gordon Amoh, Henry J. Lawson

**Affiliations:** Family Medicine/Polyclinic Department, Korle-Bu Teaching Hospital, Accra, Ghana; RHI Medical Centre, Amanokrom-Akuapem, Ghana; Department of Community and Preventive Dentistry, University of Ghana Dental School, Accra, Ghana; Department of Dietetics, School of Biomedical and Allied Health Sciences, College of Health Sciences, University of Ghana, Accra, Ghana; Department of Community Health, University of Ghana Medical School, Accra, Ghana

**Keywords:** Physical inactivity, Sedentary, Corporate bank workers, Adults, Ghana

## Abstract

**Introduction:** It is common knowledge that any activity that results in caloric expenditure has the potential to prevent cardiovascular diseases, nonetheless, most people are physically inactive, especially office workers. This study sort to evaluate at baseline, the extent of physical inactivity and its determinants among staff of selected banks in Accra, Ghana.

**Methods:** This was a cross-sectional study involving 219 banking staff randomly selected from five commercial banking institutions in Accra, Ghana. Demographic data was collected with a structured questionnaire. Physical inactivity was assessed using the Global Physical Activity Questionnaire. Study associations were determined using univariate analysis, and multivariate logistic regression models with adjusted odds ratio (AOR) and 95% confidence intervals (CI) estimated.

**Results:** Of the 219 participants studied, the male to female ratio was 1.3:1.0 and the mean age (± SD) was 40.0±7.9 years. Overall, 165 (75.3%) of the study participants indulged in some form of physical activity, however, only 40 (18.3%) achieved the recommended levels. Physical inactivity was observed in 179 (81.7%) participants. The following were independently associated with physical inactivity: travel-related activities (AOR, 0.151; 95% CI, 0.059-0.384; p<0.001); working in the bank for 6-10 years (AOR, 4.617; 95% CI, 1.590-13.405; p=0.005); and working in the bank for 11 years and above (AOR, 2.816; 95% CI, 1.076-7.368; p=0.035).

**Conclusion:** Physical inactivity was very high among bankers. Travel-related activities and working at the bank for more than six years were associated with physical inactivity. Thus, promoting regular physical activity, frequent monitoring, and implementation of other appropriate healthy lifestyle intervention strategies are vital to reduce risk of early onset disease conditions associated with physical inactivity in this population.

## Introduction

In this era of industrial revolution, activity levels have gradually declined due to new technologies which help humans perform certain activities with greater ease. Globally, about one in four adults and 81% of school-going adolescents are physically inactive [1]. Some reasons attributed to the high levels of inactivity include lack of time, financial costs, entrenched attitudes and behaviours, restrictions in the physical environment, low socioeconomic status and lack of knowledge [2]. Physical inactivity increases with age, is higher in adult women than men, and is more prevalent in high-income countries compared to low-income countries [3]. Physical inactivity is a primary but modifiable risk factor for non-communicable diseases (NCDs), particularly cardiovascular diseases (CVDs) [4–6].

Compared to other CVD risk factors, PI has been found in many studies to have the highest impact especially among employees in financial institutions [7,8]. These employees spend longer hours on their seats [9] and this is associated with high incidence of other CVD risk factors [10]. Physical inactivity is more frequently encountered in highly paid workers and in people with higher educational levels. These persons usually have fixed work programs and trade-off their leisure time with work to earn higher wages. Sedentary lifestyle, which is a behaviour conducted in sitting or reclining positions also has similar health hazards [11]. A person can be physically inactive but not necessarily sedentary. Conversely, one can be physically active but engage in high levels of sedentary behaviours.

Although people of high socioeconomic status, high educational levels and males prefer recreational-related physical activities [12,13], studies have shown that those who practice travel-related physical activities are more likely to achieve recommended physical activity levels [14,15]. Physical activity and exercise have undebatable health benefits; virtually everybody can gain from being more physically active [16]. A review of interventions (such as educational campaigns, social supports, policies etc.) across different populations, found acceptable increases in physical activity among people of various ages, social groups, communities and countries [17]. For improved health, the World Health Organization (WHO) recommends at least 150 minutes a week of moderate-intensity aerobic physical activity or 75 minutes a week of vigorous-intensity physical activity in adults [18] or an equivalent combination achieving at least 600 metabolic equivalent of task (MET). Metabolic equivalent of task is another method for characterizing physical activity at different levels of effort and its used to estimate the amount of oxygen used by the body during physical activity [19].

Physical inactivity (PI) remains one of the leading risk factors for worldwide mortality and accounts for over two thirds (2.6 million) of the mortality in low- and middle-income countries [20]. Most office work is associated with PI and sedentary lifestyle which makes them prone to diseases and reduced quality of life [8,9,21]. Although some studies have looked at physical inactivity levels in the sub-region, only few studies have looked at this working population and explored the domains of activities practiced.

The study aimed to assess the baseline PI among bankers in Accra, Ghana. The specific objectives of the study were to: estimate PI among bankers in Accra, Ghana; demonstrate the different domains of activity practiced among bankers; determine sedentary levels among bankers in Accra, Ghana and evaluate factors that are associated with PI among bankers in Accra, Ghana

## Methods

### Study design, population, and Period

This was a cross-sectional study conducted between September 2018 and February 2019. The study was conducted at the head offices of selected commercial banking institutions in the Accra Metropolis of Ghana. The head office is the centre of operations of all commercial banks and harbours all departments of the institution. Bankers constitute a category of office workers with sedentary behaviours that increase the risk of CVDs [21]. Bankers include Tellers, customer service personnel, audit and inspection officers, treasury personnel, human resource personnel, marketing personnel, accounting officers, risk and compliance officers, credit analysts, executive managers, and information technology personnel.

### Sampling, inclusion, and exclusion criteria

At the time of study, there were thirty-three (33) registered commercial banks in Ghana and thirty

(30) of them had head offices situated in the Accra metropolis [22]. From an alphabetically sorted and numbered list of all commercial banking institutions in Accra [22], five were selected as study sites using the random number generator function in Statistical Package for Social Sciences (SPSS), version 25. Using the same method, forty-five participants were randomly recruited from each selected bank. The following banking staff were excluded from the study: Pregnant women, those who joined the banking industry for the first time within 6 months before the study and those who were on routine annual leave during the study period.

### Data collection, measurements, and quality control

Data collection was done at the premises of the selected banks. A structured questionnaire which was developed from the Global Physical Activity Questionnaire was used to collect data. This consisted of open- and close-ended questions, check-list questions, and a Likert scale. The first part of the questionnaire focused on demographic information of the participants, which included gender, age, highest education level and the number of years working in the bank.

The second part measured the physical activity levels of participants across three domains: work-related (home /office) activities, travel-related activities, and recreational-related activities undertaken for at least 10-minutes continuously. Work-related activities included heavy cleaning, gardening, carrying heavy loads, construction work, digging etc. Travel-related activities included walking or using bicycles to get to and from places, while recreational-related activities included brisk walking, jogging, aerobics, cycling, soccer, lawn tennis, volleyball, etc. These were further categorized into moderate- and vigorous-intensity activities. Examples of vigorous-intensity physical activity included soccer game, fast cycling, running/jogging, hiking uphill, carrying heavy loads, heavy gardening, and aerobic dancing. Moderate-intensity physical activity also included heavy cleaning or washing, carrying light loads, general gardening, light cycling, mowing lawns, brisk walking, and tennis doubles. The total time (in minutes per week) expended in moderate and vigorous physical activity was used to sort participants into two physical activity categories as recommended by the WHO [18]. For the calculation of participants’ overall energy expenditure, the MET values for corresponding activities as specified by WHO was used: where moderate intensity activity is equivalent to MET value of 4.0 and vigorous intensity activity is equivalent to MET value of 8.0 [19].

Sedentary behaviour was assessed by the number of hours participants spend sitting or reclining on a typical working day. Sedentary behaviour was assessed as a categorical variable as high (≥8 hours per day) and normal (< 8 hours per day) [23].

### Statistical Analysis

Study data was analyzed using Statistical Package for Social Sciences (SPSS) version 25. Study outcomes were determined using uni- and bi-variate analysis, and multivariate logistic regression models. Comparisons between categorical data were done with Chi-square or Fisher’s exact test where appropriate. Continuous data were compared using student’s t-test with analyses of variance (ANOVA) for multiple comparisons. The level of statistical significance was set at p < 0.05. Socio-demographic characteristics and the domains of activity that showed significant bivariate associations (P-value <0.05) with PI were further analyzed using multivariate logistic regression to identify independent risk factors associated with physical inactivity.

### Ethical Considerations

This study received ethical clearance from the Ghana Health Service Ethics Review Committee (GHS-ERC011/05/15). Study was adequately explained to participants and consent sort before they were recruited. Permission was also sort from the management of all the banks selected for the study.

## Results

Overall, 225 professional bankers (45 each from 5 selected banks) were invited to participate in the study. Of these, 219 bankers responded and were included in the study, representing a response rate of 97.3% [95% CI: 94.3-98.7%]. The mean response rate [±standard deviation (SD)] for all selected banks was 97.3±3.65% and ranged from 91.1% to 100.0%. The male to female ratio was 1.3:1.0. The age of participants ranged from 26-60years with a mean age (±SD) of 40.0±7.9 years. Participants aged between 20-39years were 119 (54.3%) and those aged between 40-60 years were 100 (45.7%). All the participants had worked in a bank for 2-37years. Forty-five (20.5%) of the participants had worked in a bank for 1-5 years; 68 (31.1%) had worked for 6-10 years; and 106 (48.4%) had worked for more than 10years.

The prevalence of PI among the study participants was 81.7%. Altogether, 165 (75.3%) of the study participants indulged in some form of PI, either work-related, travel-related, recreational-related or a combination of these (Figure 1). Participants who practiced work-related, travel-related, and recreational-related activities were 36 (16.4%), 31 (14.2%) and 127 (58.0%) respectively. The mean metabolic equivalence of tasks (METs) per week expended by banking staff who engaged in travel-related activities (583.2) or work-related activities (487.2) was significantly higher than what was expended by banking staff who engaged in recreational-related activities (367.9); *p* < 0.05 as shown in figure 2. Overall, the mean [± SD] amount of time spent by banking staff in a sedentary lifestyle was 8.66±2.10 hours/day. About 159 (69%) of the banking staff indulged in high sedentary behaviour (Table 1)

**Fig. 1:**
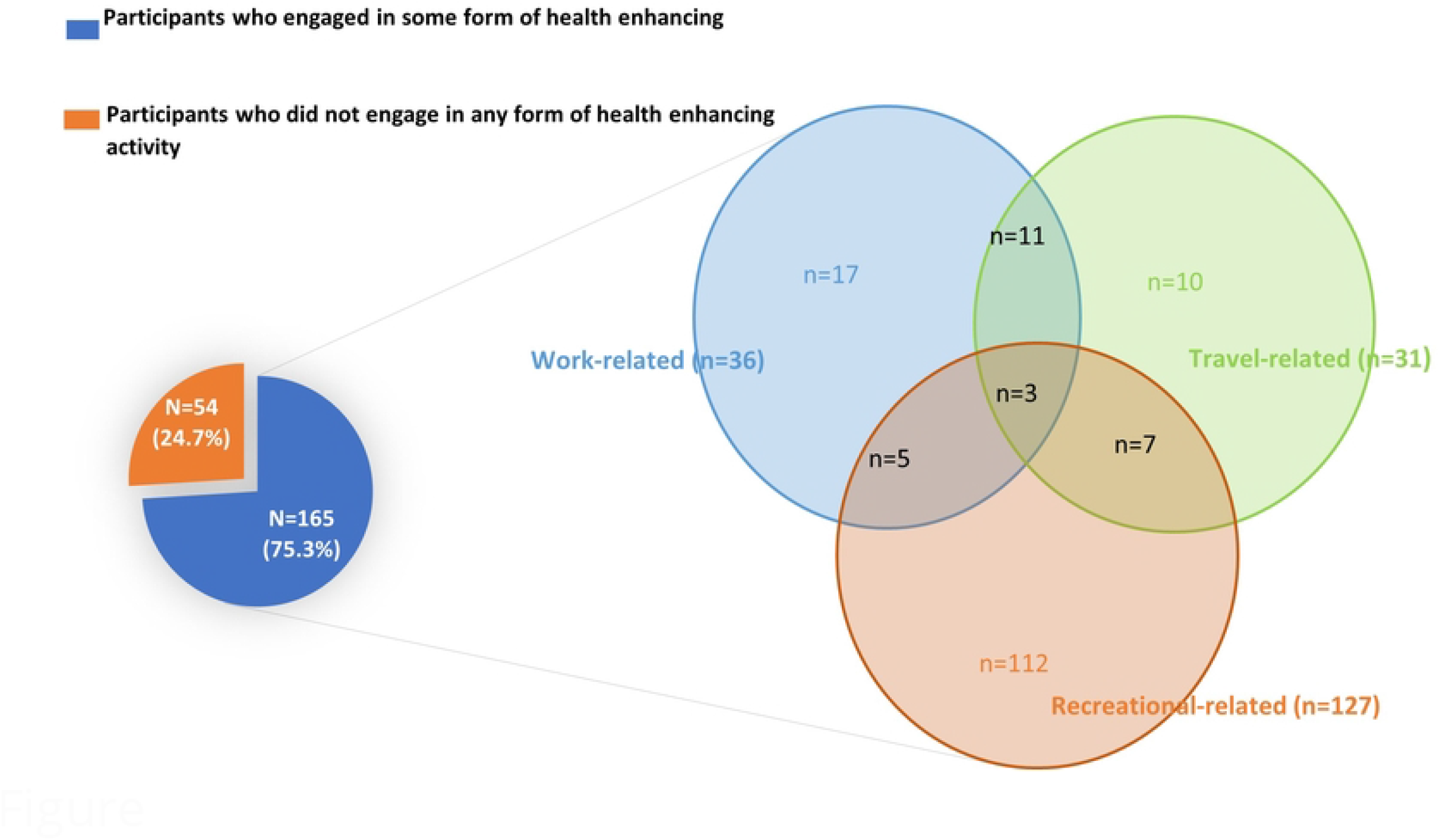
Domains of health enhancing activities. *The Venn diagram shows the various domain of activities practiced either alone or in combination as illustrated by the intersection of the sets*.

**Fig. 2:**
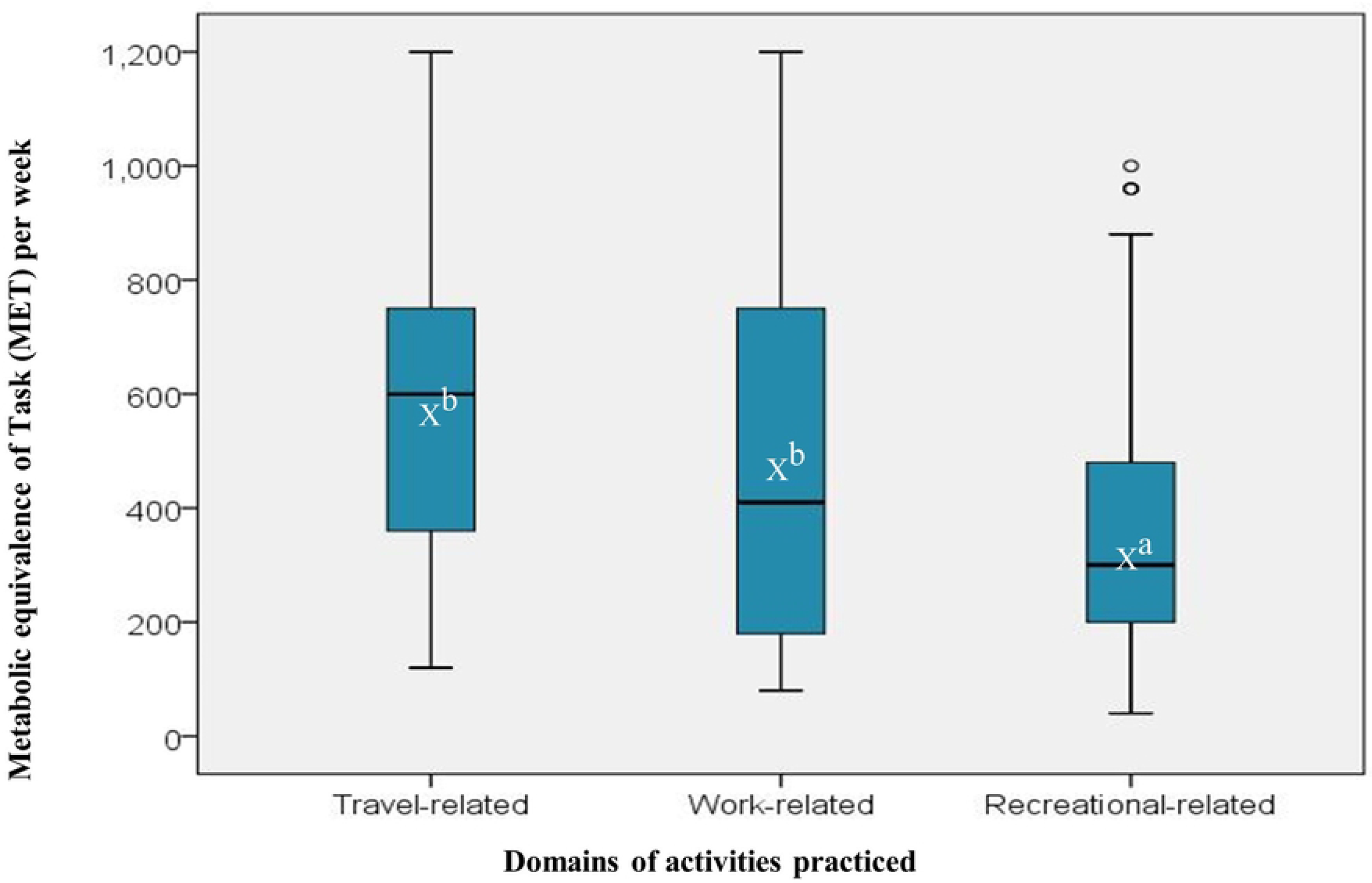
Domains of activities and metabolic equivalence tasks (METs) expended. *The box plot shows the interquartile range, horizontal line in the box shows the median of the distribution, X* denotes *mean of the distribution, lower whisker shows 25th percentile or lower quartile, upper whisker shows 75th percentile or upper quartile*. *NB. Superscripts compare the mean METs per week expended per domain of activities practiced, where b>a at P<0*.*05*.

**Table 1:** Sedentary Behaviour Among Banking Staff

|  | Normal sedentary behaviour | High sedentary behaviour | Overall |
| --- | --- | --- | --- |
| **Number of bankers** | 68 (31.1%) | 151 (68.9%) | 219<sup>a</sup> |
| **Mean time in hours per day (± standard deviation)** | 6.54 ± 0.63 | 9.64 ± 1.74 | 8.66 ± 2.10 |
| **Range** | 5.0-7.0 | 8.0-14.0 | 3.0-14.0 |
| **Interquartile range (25%, 75%)** | 6.0, 7.0 | 8.0, 11.0 | 7.0, 10.0 |

In the univariate model (Table 2), the prevalence of PI was significantly higher in participants who had worked in the bank for 6-10 years or greater than 10 years, compared to those who had worked in the bank for 1-5 years. The prevalence of PI did not differ significantly between participants who were 40-60 years compared to those between 20-39 years. The prevalence of PI did not also differ significantly between males and females nor among the various banks. The prevalence of PI was significantly lower in participants who engaged in travel- or work-related activities. There was no significant difference in the prevalence of PI among participants who engaged in recreational-related activities compared to other activities. In the logistic regression analysis using PI as the dependent outcome, the following variables were significant independent predictors of PI: working at the bank for 6-10 years (AOR, 4.617; 95% CI, 1.590 -13.405; *p*=0.005) or greater than 10 years (AOR, 2.816; 95% CI, 1.076 -7.368; *p*=0.035) and travel-related activities (AOR, 0.151; 95% CI, 0.059 - 0.384; *p*<0.001). (Table 2)

**Table 2:**
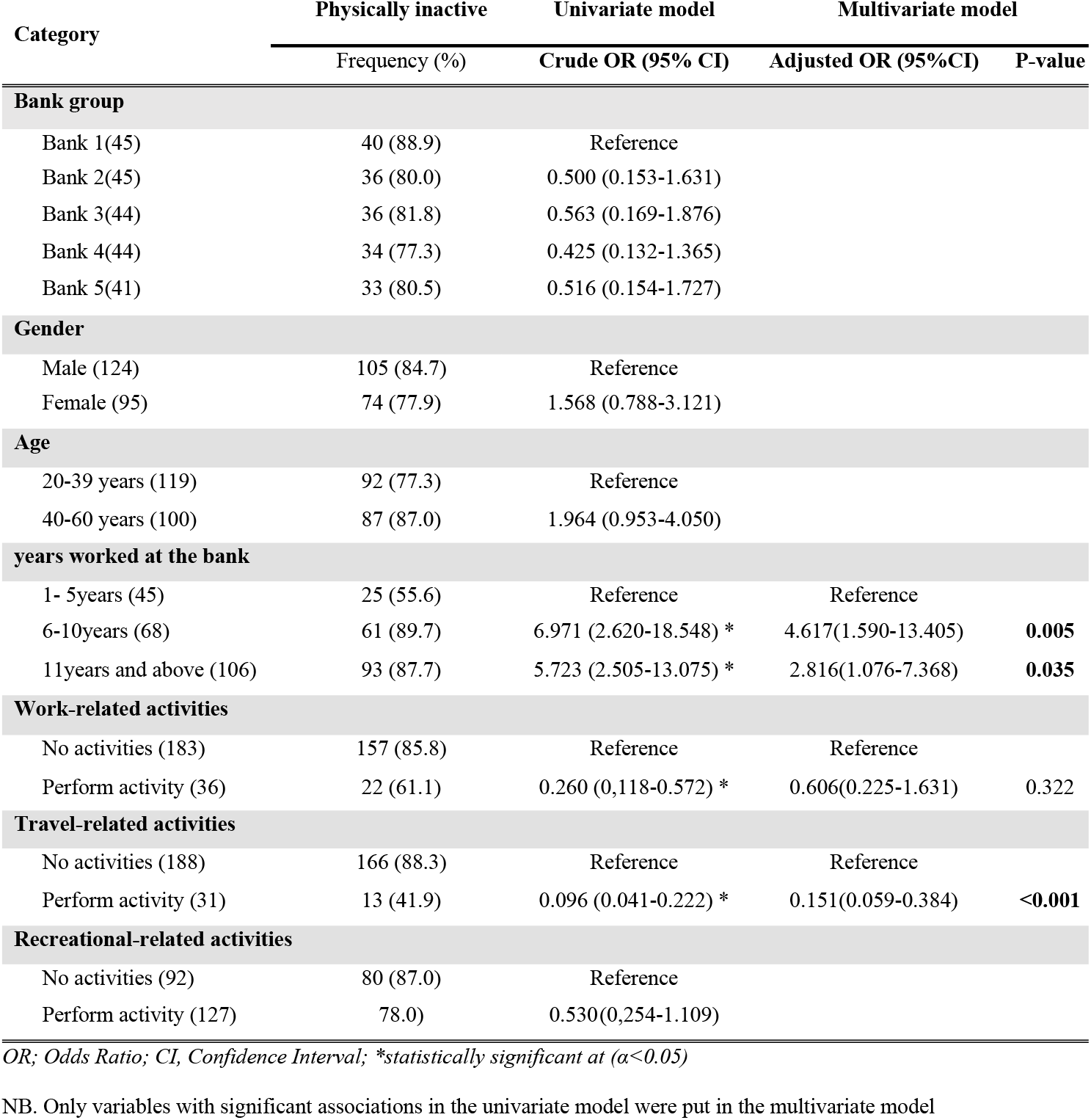
Factors Associated with Physical Inactivity

| Category | Physically inactive | Univariate model | Multivariate model |  |
| --- | --- | --- | --- | --- |
|  | Frequency (%) | Crude OR (95% CI) | Adjusted OR (95%CI) | P-value |
| **Bank group** |  |  |  |  |
| Bank 1(45) | 40 (88.9) | Reference |  |  |
| Bank 2(45) | 36 (80.0) | 0.500 (0.153-1.631) |  |  |
| Bank 3(44) | 36 (81.8) | 0.563 (0.169-1.876) |  |  |
| Bank 4(44) | 34 (77.3) | 0.425 (0.132-1.365) |  |  |
| Bank 5(41) | 33 (80.5) | 0.516 (0.154-1.727) |  |  |
| **Gender** |  |  |  |  |
| Male (124) | 105 (84.7) | Reference |  |  |
| Female (95) | 74 (77.9) | 1.568 (0.788-3.121) |  |  |
| **Age** |  |  |  |  |
| 20-39 years (119) | 92 (77.3) | Reference |  |  |
| 40-60 years (100) | 87 (87.0) | 1.964 (0.953-4.050) |  |  |
| **years worked at the bank** |  |  |  |  |
| 1- 5years (45) | 25 (55.6) | Reference | Reference |  |
| 6-10years (68) | 61 (89.7) | 6.971 (2.620-18.548) * | 4.617(1.590-13.405) | **0.005** |
| 11years and above (106) | 93 (87.7) | 5.723 (2.505-13.075) * | 2.816(1.076-7.368) | **0.035** |
| **Work-related activities** |  |  |  |  |
| No activities (183) | 157 (85.8) | Reference | Reference |  |
| Perform activity (36) | 22 (61.1) | 0.260 (0.118-0.572) * | 0.606(0.225-1.631) | 0.322 |
| **Travel-related activities** |  |  |  |  |
| No activities (188) | 166 (88.3) | Reference | Reference |  |
| Perform activity (31) | 13 (41.9) | 0.096 (0.041-0.222) * | 0.151(0.059-0.384) | **<0.001** |
| **Recreational-related activities** |  |  |  |  |
| No activities (92) | 80 (87.0) | Reference |  |  |
| Perform activity (127) | 78.0) | 0.530(0.254-1.109) |  |  |

OR; Odds Ratio; CI, Confidence Interval; \*statistically significant at ( $\alpha < 0.05$ )
NB. Only variables with significant associations in the univariate model were put in the multivariate model

## Discussion

Physical inactivity continues to saddle society with a hidden and growing cost of healthcare and loss of productivity [1]. This study assessed PI among banking staff in Accra. The age range of the study population (26-60years) was characteristic of the typical working-age group in Ghana [24]. All the participants had spent at least 15 years of formal education after preschool which presupposes that most participants had received tertiary education.

The study reported a very high prevalence of PI (81.7%) among banking staff compared to the prevalence of 21.7% estimated for the general Ghanaian population from the RODAM study [25]. This contrast may be due to the stressful nature of the banking profession coupled with their long working hours [26]. This lifestyle denies bankers adequate time for workouts. In a recent report on the barriers to the uptake of healthy behaviours, Kelly *et al* [2] described inadequate time for exercise as a major reason for recent low levels of physical activity. The high level of PI observed in this study is worrying, because of its strong association with high cardiovascular mortality [27,28] however, it agrees with the 83.3% prevalence reported among workers in some financial institutions in Accra in 2015 [8]. In a similar study among bankers in Nigeria, Aderibigbe and colleagues observed a 60% prevalence of PI which is lower than what was recorded in the current study [29]. It is important to note that the prevalence of PI in this study was comparable across the five banks studied. This observation is in keeping with data from global reviews and across Ghana in particular which suggest that working conditions that predispose staff to sedentary lifestyle and PI are similar in most banks [26,30]. It’s noteworthy that, 57 of the 179 physically inactive banking staff did not indulge in any form of activity lasting at least 10minutes. The remaining 122 bankers practiced some form of activity, nevertheless, these activities were not enough to meet the recommended level to be considered physically active. This observation may be attributable to the inadequate time available to banking staff for exercise

Overall, a total of 165 study participants indulged in some form of activity lasting at least 10 minutes. The three domains of activities practiced by bankers were recreational-, work-, and travel-related activities. Recreational-related activities were the predominant (58%) domain of activity practiced by the bankers. This observation somewhat supports the finding by Nang *et al*. who reported that recreational-related activities are often the mainstay of persons within the higher socioeconomic bracket [13]. Beenackers *et al*. also reported in their systematic review that people of a high socioeconomic class are more likely to engage in recreational-related activities than people of low socioeconomic class because of resource availability and access to recreational facilities [31]. This may be the case of banking staff as most of them have access to recreational facilities. Fewer participants indulged in significant work- or travel-related activities. Most bankers belong to the high socioeconomic group of workers and are likely to have personal cars or house-helps which may reduce their travel-related and/or work-related activities at home. The sedentary nature of their work does not support work-related activity at the workplace. This supports the hypothesis of a changing social pattern of physical activity, with a decline in occupational physical activity, especially among the workers in the formal sector [32]. The use of personal cars as modes of transport, house-helps, and workplace technologies have been found in the study by Owen *et al*. to reduce work- and travel-related activities, especially among persons of high socioeconomic status [33]. In this study, banking staff who practiced travel- and work-related activities were observed to significantly have a lower prevalence of PI compared to those who did not practice such activities. In contrast, no significant difference in the prevalence of PI was observed between banking staff who practiced recreational-related activities and those who did not.

In another observation, the total energy (in METs) expended by bankers who practiced either travel- or work-related activities was significantly higher compared to those who practiced recreational-related activities. This result is in line with data from Nang *et al*. who demonstrated that although some people prefer recreational-related activities, they achieve less total activity to reach the expected minimum of 600 METs per week [13]. Engaging in work- or travel-related activities on the other hand, contributes substantially more to, and is associated with a commensurate increase in total physical activity [13,15]. Travel- and work-related activities are generally more routine and often involve longer durations. This allows those who practice it better chances of achieving recommended daily levels of physical activity. Rather than focusing on one domain of physical activity, engaging in activities across multiple domains may further augment the total physical activity achieved. Although WHO recommends physical activity across all domains to achieve the recommended activity levels, only 26 of the participants in this study indulged in two (2) or more domains of activity. Nang *et al*. also demonstrated in their study that engaging in multiple domains of activity increases the chances of achieving higher physical activity [13]. This may explain why the prevalence of inactivity was high among the banking staff although majority engaged in recreational-related activities.

Nearly 70% of the participants indulged in highly sedentary behaviour. This prevalence is considerably higher than that reported in the general population (8.3%) for low and middle-income countries [34]. Over the past decade, there is increasing literature to demonstrate a strong morbidity risk associated with a sedentary lifestyle, with some studies putting the sedentary lifestyle at the same level as smoking [35–37]. The average estimated time spent in sedentary behaviour among the participants was 8.66 hours per day. This result, coupled with the high level of PI among participants, it is safe to suggest that working in banks predisposes individuals to very high health risks.

In this study, travel-related activities and working in the bank for more than 6years were independent predictors of PI. Whiles travel-related activities reduced the risk of being inactive by about 85%, working at the bank for more than 6years increased the risk of being physically inactive by between three to five-fold, with the risk being lower as staff remain in the bank for more than 10years. The findings are consistent with a study by Knell *et al* [38], who concluded that travel-related activities were associated with high predictability (AOR, 7.3: 95% CI, 2.6-20.1) of meeting the physical activity recommendation. Salquist *et al*.[15] also concluded in their study that, substantial physical activity can be accumulated through travel-related activities. Travel-related activities generally involve longer durations of time and can help achieve the recommended levels of physical activity. Travel-related activities should, therefore, be encouraged to promote physical activity. A realization of the sedentary nature of the banking work and the possible health risks it poses may have prompted staff who had worked for longer years to do something about their activity. This could explain the reduction in the odds of being inactive among staff who had worked at the bank for longer years. Further studies may however be needed to authenticate this submission.

Some workers have identified other independent predictor variables for PI. In the study by Mengesha *et al*., being a female adult and having office employment in Ethiopia were significantly associated with PI[39]. In Kenya, Gichu *et al*. also reported that female gender and age 30-49 years were independent predictors of PI [40]. This study however did not find female gender or age to be an independent predictor of physical inactivity.

Based on the findings of this study, we recommended that banking staff are encouraged to add travel-related activities to their domain of activities practiced because the results show that bankers who practiced travel-related activities achieved higher energy in MET per week. Travel-related activities independently decreased the likelihood of being physically inactive among the banking participants. Travel-related activities could, therefore, help bankers meet their recommended physical activity levels for health. Additionally, comprehensive workplace health promotion is recommended for all banking institutions to help improve healthy behaviours, particularly physical activity. Since bankers spend most of their wakeful time at the workplace, worksite health programs that promote physical activities will go a long way to help.

The study had some limitations that are worth noting. Data collected on participant’s activity levels were based on self-report and thus liable to recall bias. Secondly, the study did not discriminate participants by department in the bank because the design and functions of some departments make it unavoidable for staff to be sedentary for long periods.

## Conclusion

The study reported high levels of PI among bankers, with majority indulged in high sedentary behaviours. The predominant domain of activity practiced by the participants was recreational-related although those who engaged in either travel-related or work-related activities significantly expended higher energy in METs per week compared to those who practiced recreational-related activities. Engaging in travel-related activities decreases the likelihood of being inactive. Then again, working at the bank for more than six years increases the likelihood of being inactive. Hence there is the need for a concerted effort to encourage regular physical activity both outside and around the working environment and to implement other healthy lifestyle practices, as well as the continued monitoring of these behaviours, to prevent early onset of non-communicable diseases among this population.

## Data Availability

It will be made available on request

## Acknowledgement

We thank MDS-Lancet Laboratories for their support, and to all the respondent banks and their staff who consented to make this study possible.

## Authors Contribution

**Conceptualization**: George B. Nketiah, Kwasi Odoi-Agyarko, Henry J. Lawson

**Data Curation:** George B. Nketiah, Tom A Ndanu, Gordon Amoh, Frank E A Hayford

**Formal Analysis**: George B. Nketiah, Tom A Ndanu, Kwasi Odoi-Agyarko

**Investigation:** George B. Nketiah, Gordon Amoh

**Methodology**: George B. Nketiah, Kwasi Odoi-Agyarko, Tom A Ndanu, Gordon Amoh, Frank E A Hayford, Henry J. Lawson

**Project Administration:** George B. Nketiah

**Resources:** George B. Nketiah, Kwasi Odoi-Agyarko

**Software:** George B. Nketiah, Tom A Ndanu

**Supervision:** Kwasi Odoi-Agyarko, Henry J. Lawson

**Validation:** Kwasi Odoi-Agyarko

**Visualization:** Henry J. Lawson

**Writing** – Original Draft Preparation: George B. Nketiah, Gordon Amoh

**Writing** – Review & Editing:Kwasi Odoi-Agyarko, Henry J. Lawson, Tom A Ndanu, Frank E A Hayford

All authors approved final draft of the manuscript before submission.

